# Time of surgery for aneurysmal subarachnoid hemorrhage in patients ≥70 years

**DOI:** 10.1101/2023.12.12.23299889

**Authors:** Hengrui Zhang, Bangyue Wang, Ruyi Wang, Chao Peng, Changkai Hou, Yan Zhao, Linchun Huan, Yanfen Chai, Xinyu Yang, Jianjun Yu

**Affiliations:** Department of Neurosurgery, Tianjin Medical University General Hospital, Tianjin, China; Department of Emergency Medicine, Tianjin Medical University General Hospital, Tianjin, China; Department of Neurosurgery, Linyi People’s Hospital, Linyi, China

**Author notes:** **Corresponding author** Jianjun Yu M.D. Department of Neurosurgery, Linyi People’s Hospital, No. 27 Jie-Fang Road, Linyi 276000, China. Joint first authors (Hengrui Zhang, Bangyue Wang, Ruyi Wang). Joint senior authors (Yanfen Chai, Xinyu Yang, Jianjun Yu).

## Abstract

**Objective:** To establish a time-to-surgery threshold for elderly aneurysmal subarachnoid hemorrhage patients before the risk of mortality increases.

**Methods:** A cohort study using data with consecutive patients 70 years and older(N=743). Risk-adjusted restricted cubic splines modeled the mortality according to wait-time. The inflection point (in hours) when mortality began to increase was used to define early-time, middle-time and late-time surgery. To evaluate the robustness of this definition, outcomes among propensity-score matched non-middle surgical and middle-time surgical patients were compared using percent absolute risk differences.

**Results:** There were a total of 535 patients who met inclusion. Their mean age was 74.3 (4.38) years. Cox models with restricted cubic spline showed a statistically significant U shaped association for onset-to-surgery time with two year all cause mortality. 66 patients (12.3%) received middle-time surgery and 469 patients (87.7%) received early-time or late-time surgery. There were 60 (91%) patients in the middle-time surgery group matched to those in the non-middle-time group. Of the 60 matched patients who received surgery after less than 49 hours or more than 68 hours, 23 patients (38.3%) died within 24 months vs 14 patients (23.3%) of 60 who received surgery within 49 to 68 hours, for an absolute risk difference of 15% (95% CI, -2.68% to 31.50%).

**Conclusions:** In this study, onset-to-treatment time showed a U shaped association with 24 months all cause mortality. Early surgery was superior to delayed surgery in reducing death rate. Elderly patients with poor tolerance to ultra-early surgery in whom operation was probably postponed.

## Introduction

The incidence of spontaneous aneurysmal subarachnoid hemorrhage (aSAH) does not decrease and probably even increases after the seventh decade of life.^1^ Elderly aSAH patients are characterized by high early mortality.^2^ It was reported that the high mortality and morbidity in an early surgery group was attributed to brain edema, vasospasm and poor tolerance to the surgery.^3^ Most likely because of better diagnosis, early aneurysm repair, and advanced intensive care support, the timing of treatment for aSAH has experienced a shift from favoring delayed treatment in the 1970s to preferring early treatment in recent years.^4–7^

Controversy persists regarding the acceptable waiting time for treatment. Clinical guidelines recommend treatment as early as feasible ^8^ or within 3 days after the onset of symptoms.^9^ In some studies, the optimal timing of treatment is 1 or 2 days for aSAH.^3,10,11^ The inconsistent results highlight the lack of evidence for the best timing of treatment in elderly aSAH patients.

Most previous studies among patients with aSAH were based on small samples and were not restricted to older people,^12^ or wait times were measured imprecisely in days and arbitrarily divided into early and delayed groups,^3,5,10^ decreasing the statistical power to find differences. Furthermore, the correlation between onset-to-treatment time and the outcome may be nonlinear. To fill this data gap, the present study specifically focused on the subgroup of elderly aSAH patients (age ≥70 years). The objective of this study was to establish a time-to-surgery threshold for elderly aSAH patients before the risk of mortality increases.

## Methods

### Study design and setting

We performed a cohort study using prospectively collected data from the Chinese Multicenter Cerebral Aneurysm Database (CMAD), which involves 12 comprehensive cerebrovascular centers in northern China. The CMAD was designed as a multicenter, prospective cohort study of aneurysmal SAH in patients in the Chinese population and registered with the Chinese Clinical Trial Registry (registration number ChiCTR2100054014). Approval for data collection was obtained from the institutional research ethics board of all participating centers. Patient consent was not required due to the retrospective nature of this study. This study followed the Strengthening the Reporting of Observational Studies in Epidemiology (STROBE)^13^ reporting guidelines(eAppendix G in the Supplement).

### Patients

We included consecutive patients 70 years and older (N=743) in the CMAD between January 1, 2017, and December 31, 2020. All patients who visited a cerebrovascular center during the enrollment period and met the inclusion criteria were asked to join this study.

The inclusion criteria were as follows: (1) age ≥70 years with saccular ruptured intracranial aneurysms; (2) SAH diagnosed by head computed tomography (CT) and/or lumbar puncture; (3) diagnosis of intracranial aneurysms confirmed by at least one examination through computed tomography angiography (CTA), digital subtraction angiography (DSA) or magnetic resonance angiography (MRA); and (4) having undergone surgical treatment, including neurosurgical and endovascular treatment, during hospitalization at the involved medical centers.

The exclusion criteria were as follows: (1) the onset time, admission time and operation time could not be determined; (2) patients who had had previous episodes of intracranial hemorrhage of unknown or untreated cause; (3) initial negative angiogram; and (4) multiple aneurysms in which the source of bleeding was unclear.

### Main Exposure

The primary independent variable was the onset-to-surgery time, which was calculated as the difference in hours between the date and time of onset of aSAH symptoms (recorded from ambulance notes and the emergency department) and the date and time of surgery (determined from procedure notes). Timing of surgery was measured from admission time to surgery time within a single surgical hospitalization record (eAppendix A in the Supplement). We captured the time that elapsed during transfer between hospitals.

### Covariates

Patient characteristics previously shown to explain most of the variation that affects the prognosis of aneurysmal SAH and time to treatment were measured, including age, sex, and comorbidities (Table 1).^3^ The type of admission (direct admission vs. transfer), hospital teaching status ^11^ (teaching vs. nonteaching hospital) and time of admission (weekend admission vs. weekday admission) are prehospital factors that influence treatment time. Teaching hospitals may be able to provide early treatment because of the early mobilization of hospital resources and the readily available staff to attend to the patient’s care needs in an emergency. Weekday admissions appear to be associated with early treatment. Associated conditions at the time of hospital presentation were also assessed: the World Federation of Neurosurgical Societies (WFNS) score^14^ and the modified Fisher scale.^15^ Poor clinical condition on admission was defined as a class III-V Hunt and Hess classification (HH) and a IV-V score on the World Federation of Neurological Surgeons (WFNS) scale. Additional covariates included treatment modality (neurosurgical or endovascular treatment) and aneurysm location (categorized into four traditional groups: internal carotid artery (ICA), middle cerebral artery (MCA), anterior cerebral artery (ACA) and posterior circulation aneurysms (PCircAs)).

**Table 1.**
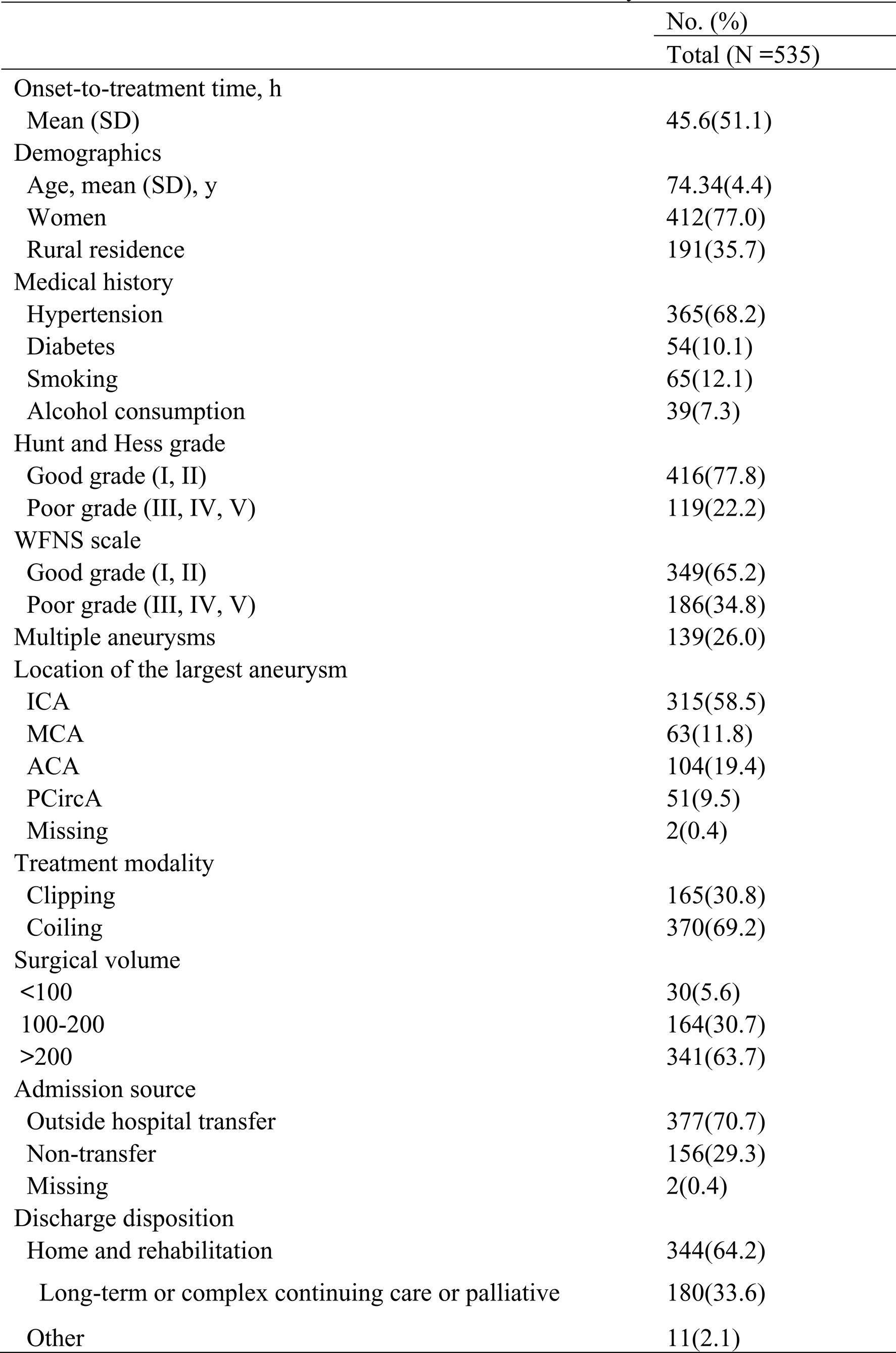
Characteristics of 535 Patients Included in the Analyses

### Outcome

The primary outcome was mortality within 2 years of being admitted for intracranial aneurysm surgery. Medical complications within 6 months, 12 months, and 24 months, as well as a composite of mortality, were assessed as secondary outcomes.

### Statistical Analysis

Descriptive statistics are displayed as the means with standard deviations (SDs) for continuous variables and as numbers (percentages) for categorical variables. For nonnormally distributed continuous variables, the medians with interquartile ranges (IQRs) are reported. Restricted cubic splines with 3 knots^16^ were used to model the probability of mortality according to the time elapsed from onset of aSAH symptoms to surgery, which generated nonlinear models and made no assumption regarding the shape of the function.

We used restricted cubic spline models fitted for Cox proportional hazards models, and a hazard ratio of 1 was the reference value. The association between surgical delay and mortality was graphically represented to visualize an inflection point at which time the risk of mortality increased significantly. Accordingly, considering onset-to-surgery time as a continuous variable, we stratified the participants into two categories (early-time or late-time, and middle-time) based on the Cox models with cubic spline models (time<49 or =70 hours and 49-70 hours). ^17^ Rather than arbitrarily dividing patients into early-time and late-time surgery groups, to further evaluate the robustness of this definition, early-time or late-time and middle-time groups were matched 1:1 without replacement^18^ on the logit of propensity scores. McNemar’s test compared outcomes in the middle and nonmiddle surgical groups after matching. Restricted cubic spline and propensity score models were all adjusted for the same covariates: sex, rural residence, hypertension, diabetes, smoking, alcohol consumption, treatment modality, multiple aneurysms, admission source, clinical condition on admission, admission source, and discharge disposition.

The threshold for statistical significance was a 2-sided *P* <0.05. Statistical analyses were performed using IBM SPSS Statistics for Windows version 27.0 (IBM Corp., Armonk, NY, USA) and R version 4.2.1 (R Core Team, Vienna, Austria).

### Sensitivity Analyses

Several prespecified sensitivity analyses were conducted. The time of surgery determined from procedure notes was likely to be highly accurate. However, the potentially least accurate time may have been the patient-reported date and time of aSAH symptom onset, which was captured from records from ambulance notes and the emergency department. To capture the time that elapsed during transfer between hospitals, all contiguous hospitalizations from the earliest hospital arrival time to the date and time of surgery were assessed. Despite all this, there was an error in the prehospitalization time, which is likely to have affected our results. To examine this, we repeated our analyses on the time to treatment excluding the prehospital time. Therefore, we used restricted cubic spline models to model the probability of mortality according to the time elapsed from first hospital arrival to surgery (eAppendix C in the Supplement).

Outside hospital transfer and poor clinical condition on admission may prolong wait times and influence outcomes.^19–21^ Therefore, an analysis was conducted to assess the association between wait times and characteristics of patients (i.e., admission source and clinical condition on admission) in a subgroup to verify our results. Poor clinical condition on admission was defined as a class III-V Hunt and Hess classification (HH) and a IV-V score on the World Federation of Neurological Surgeons (WFNS) scale. Splines were also generated in patient subgroups stratified by sex, treatment modality, and residence to determine whether the identified time threshold was consistent across patient subgroups.

Post hoc sensitivity analysis was also conducted. As an indicator of residual confounding in the early-time or late-time group, the relationship between matching status (matched or unmatched) and mortality in early-time or late-time group patients was also explored (eAppendix F in the Supplement).

## Results

### Baseline Characteristics and Sample Size

There were 535 patients who met the inclusion criteria (Figure 1) and were treated by neurosurgeons at 12 hospitals. Their mean (SD) age was 74.3 (4.38) years, and 77.0% were women. The onset time, admission time or operation time were missing for 52 patients. The mean (SD) time to surgery was 45.6 (51.1) hours.

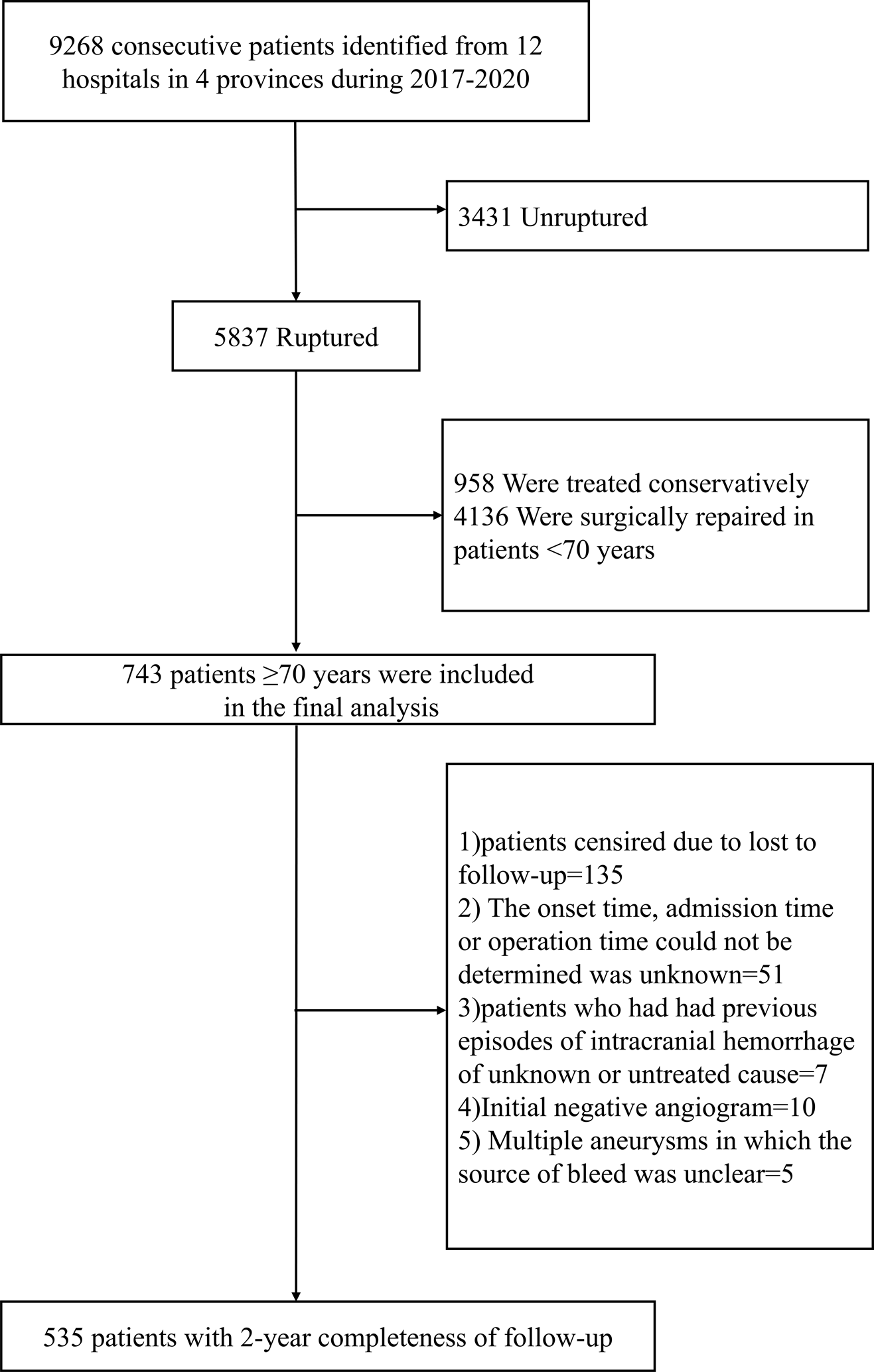

### Association of onset-to-surgery time with all-cause mortality

Cox models with restricted cubic splines showed a statistically significant U-shaped association for onset-to-surgery time with two-year all-cause mortality. Participants with treatment at approximately 68 hours after onset had the minimum mortality risk based on the estimated parameters of restricted cubic splines after adjustment (Figure 2). Compared with individuals who received treatment at approximately 68 hours after onset, the risk of all-cause mortality at two years was significantly higher among participants who received treatment at approximately less than 49 hours or more than 68 hours after onset (P=0.003, Figure 2). Inappropriate timing of surgery was therefore defined as surgery occurring less than 49 hours or more than 68 hours after onset.

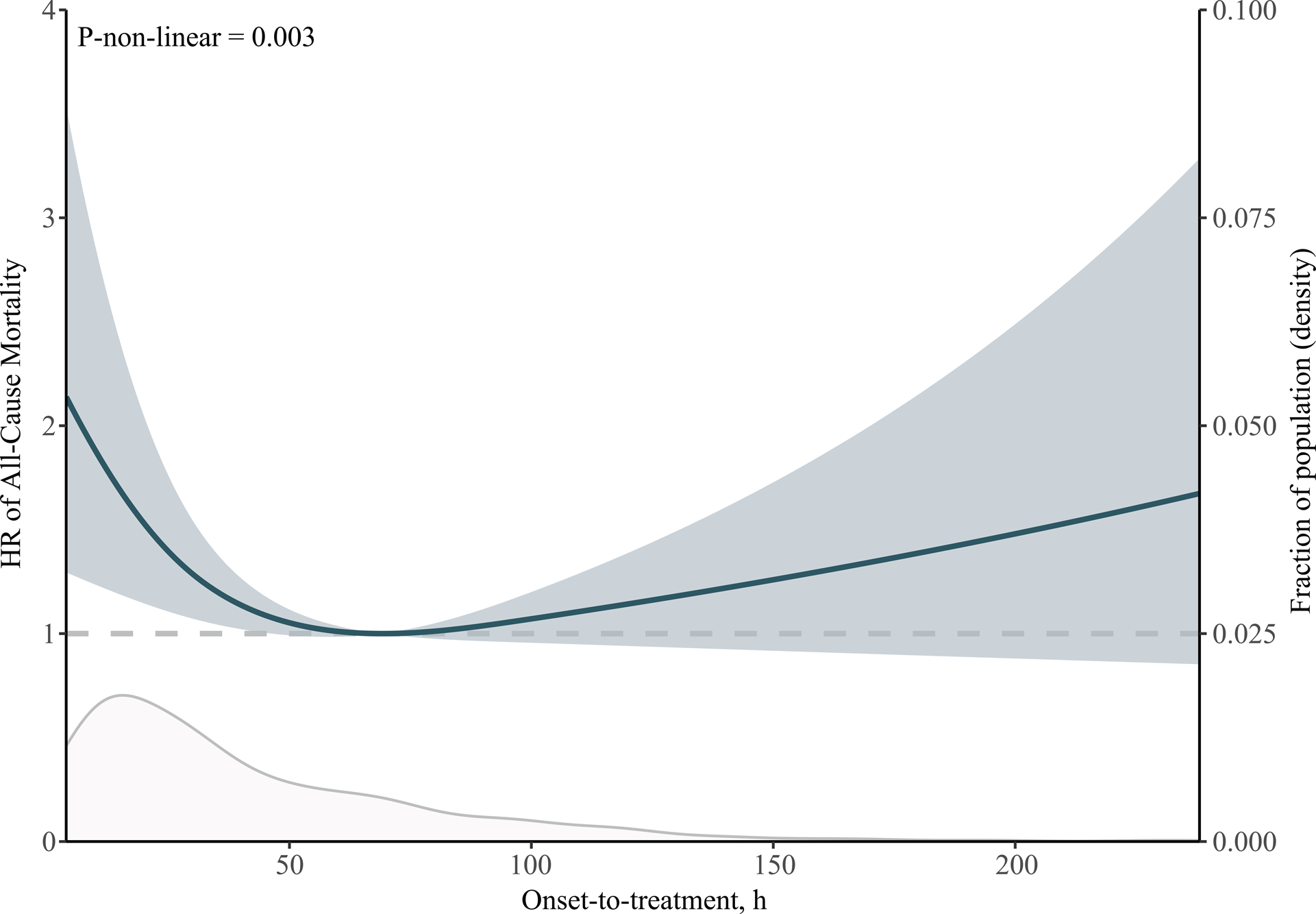

According to this definition, 66 patients (12.3%) received middle-time surgery, and 469 patients (87.7%) received early-time or late-time surgery. Before matching, patients receiving early-time or late-time surgery were significantly more likely to have poor clinical conditions on admission. Patients receiving middle-time surgery were significantly more likely to arrive from other hospitals (Table 2). There were 60 (91%) patients in the middle-time surgery group matched to those in the early-time or late-time group, and covariates were balanced between groups after matching. Of the 60 matched patients who underwent surgery at less than 49 hours or more than 68 hours, 23 patients (38.3%) died within 24 months vs. 14 patients (23.3%) of 60 who underwent surgery within 49 to 68 hours, for an absolute risk difference of 15% (95% CI, -2.68% to 31.50%); 19 (31.7) vs. 12 (20.0) died within 12 months, for an absolute risk difference of 11.67% (95% CI, -5.18% to 27.72%); and 18 (30.0) vs. 10 (16.7) died within 6 months for an absolute risk difference of 13.33% (95% CI, -3.04% to 28.90%). The rate of rebleeding within 24 months in the middle-time surgery group was 1.8% compared with 3.6% in the early-time or late-time surgery group, for an absolute risk difference of 1.75% (95% CI, -7.89% to 11.71%) (Table 3).

**Table 2.**
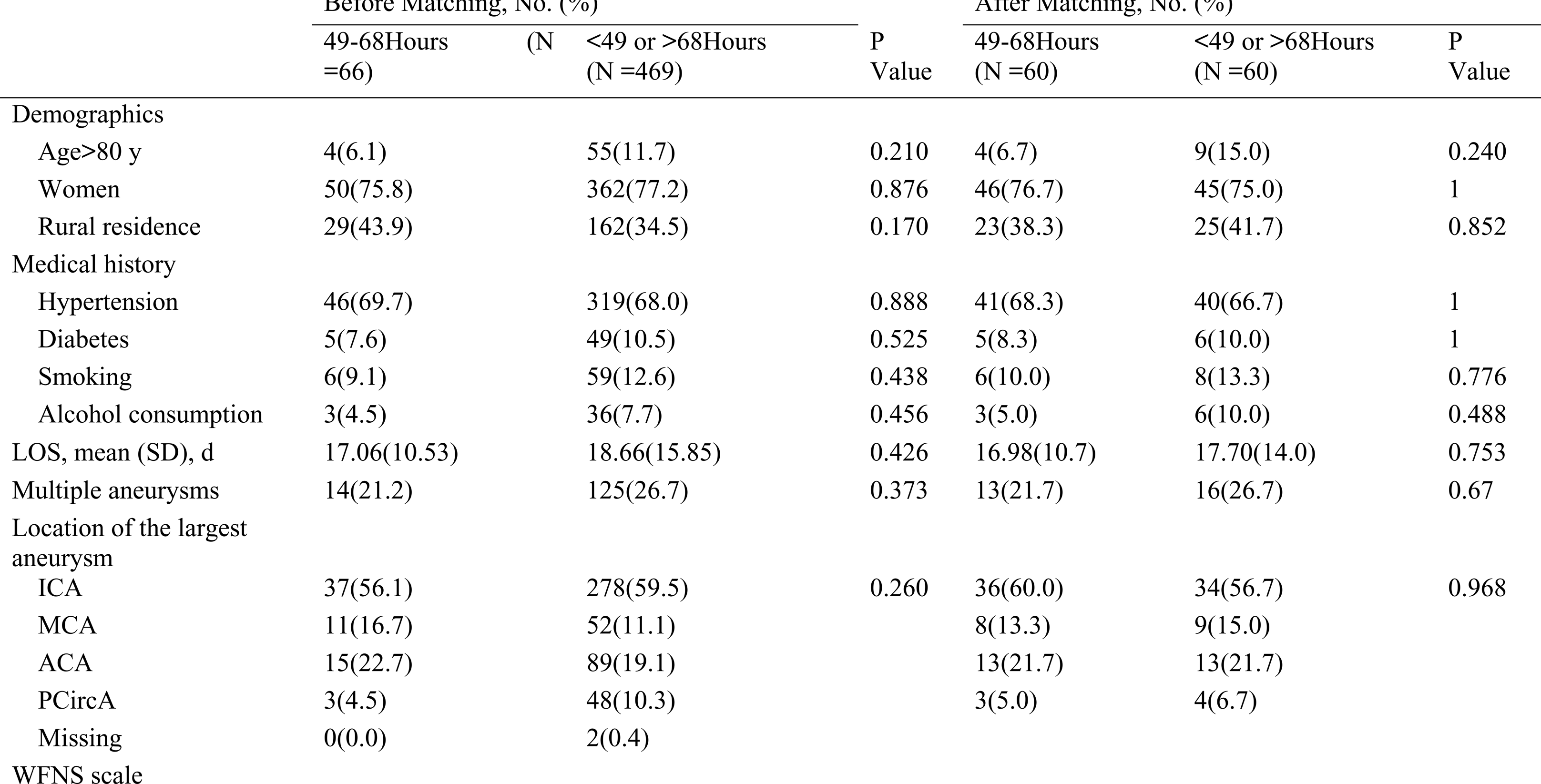
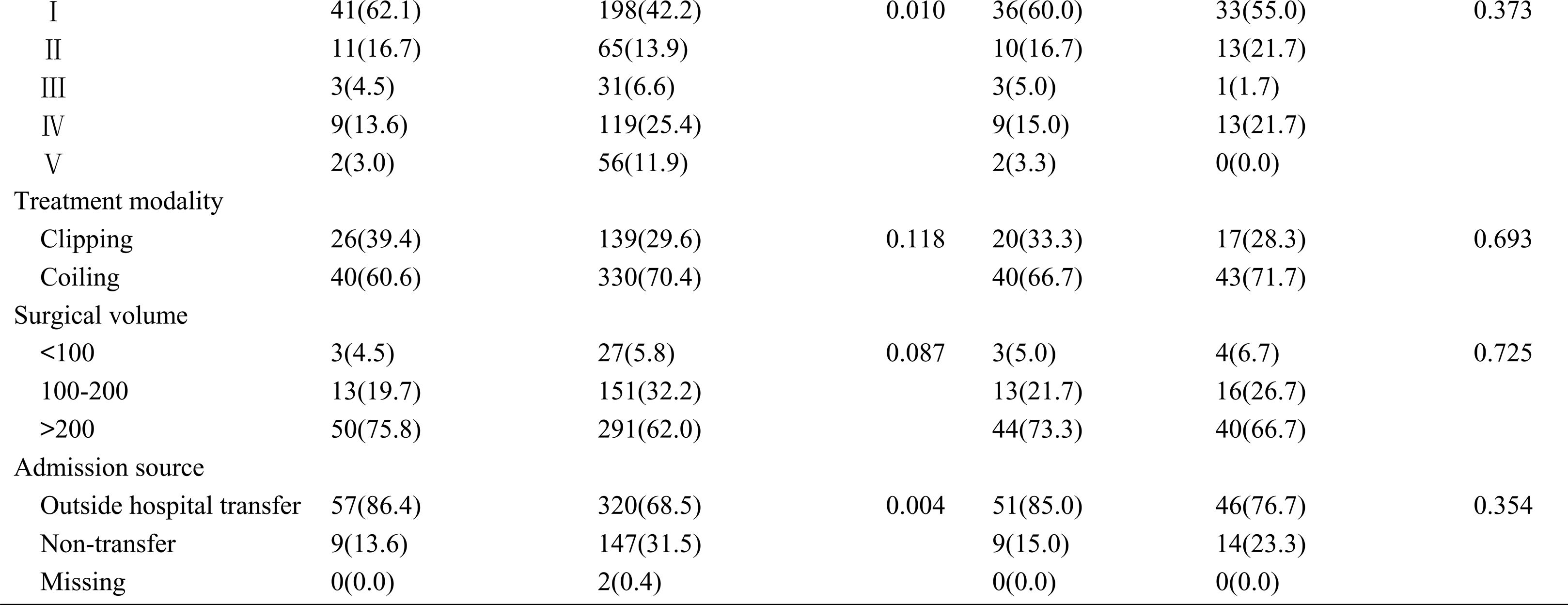
Baseline Characteristics of 535 patients undergoing intracranial aneurysm surgery in northern Chinese provincial regions between 2017-2020.

**Table 3.**
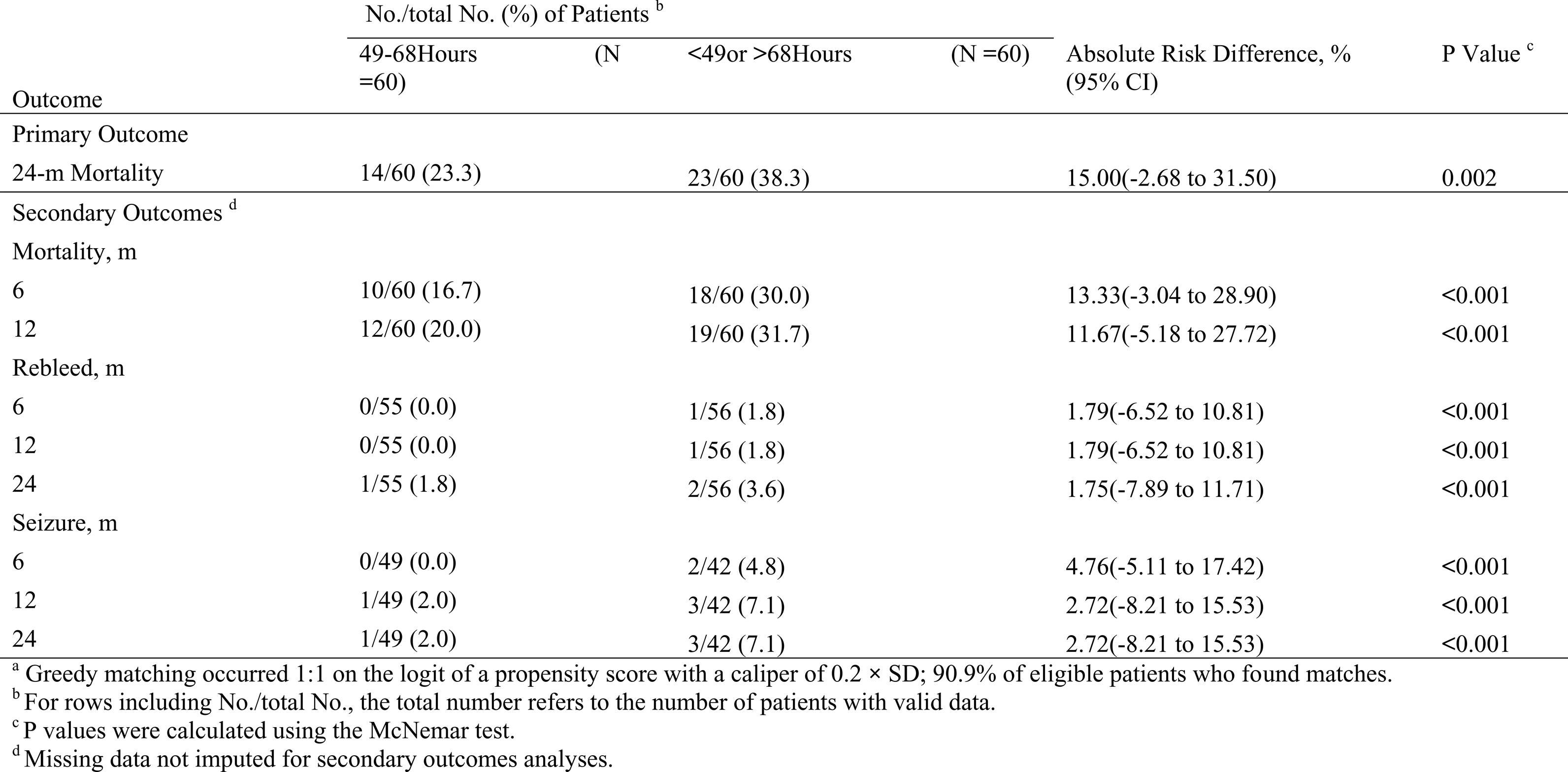
Primary and Secondary Outcomes After Matching. ^a^

Sensitivity analysis to test the accuracy of the prehospitalization time. We repeated our analyses on the time to treatment, defined as the time between the first hospital arrival and the time at which treatment was received (in hours). We found that the results were similar to those of the main analyses. Participants with treatment at approximately 68 hours after first hospital arrival had the minimum mortality risk based on the estimated parameters of restricted cubic splines after adjustment. (eAppendix C in the Supplement). The distribution of the time variable in the two models was very similar. The Bland‒Altman plot also suggested very high agreement between these two time measures (eAppendix C in the Supplement).

### Association of onset-to-surgery time with all-cause mortality in subgroups

These stratified analyses were conducted to determine whether the time threshold identified was consistent between different patient subgroups. Transfer and poor health on admission may prolong wait times. We included only those transferring and found that the onset-to-surgery time did not extend (eAppendix D in the Supplement). For patients with poor clinical condition on admission, there was a nonlinear effect even in the adjusted models. The characteristics of patients with aSAH according to condition on admission are shown in eAppendix E in the Supplement. Patients in poor health on admission were significantly more likely to have shorter wait times (P<0.001), transfer from outside the hospital (P<0.001), and have long-term or complex continuing care or palliative care (P<0.001). We found that 48.6% of patients (n =107) with poor health on admission died at 24 months compared with 16.5% (n =52) with good health on admission (P <0.001). The characteristics of patients with aSAH according to residence are shown in eAppendix E in the Supplement. Patients who lived in the city were significantly more likely to have multiple aneurysms (P=0.029), coil intracranial aneurysms (P=0.033), and transfer from outside hospitals (P<0.001). Splines were also generated in patient subgroups stratified by sex, treatment modality, and residence (Figure 3). We found that the identified time threshold was consistent across patient subgroups. Significant U-shaped associations of onset-to-treatment time with 24-month all-cause mortality were evident for many subgroups but not for clinical condition on admission.

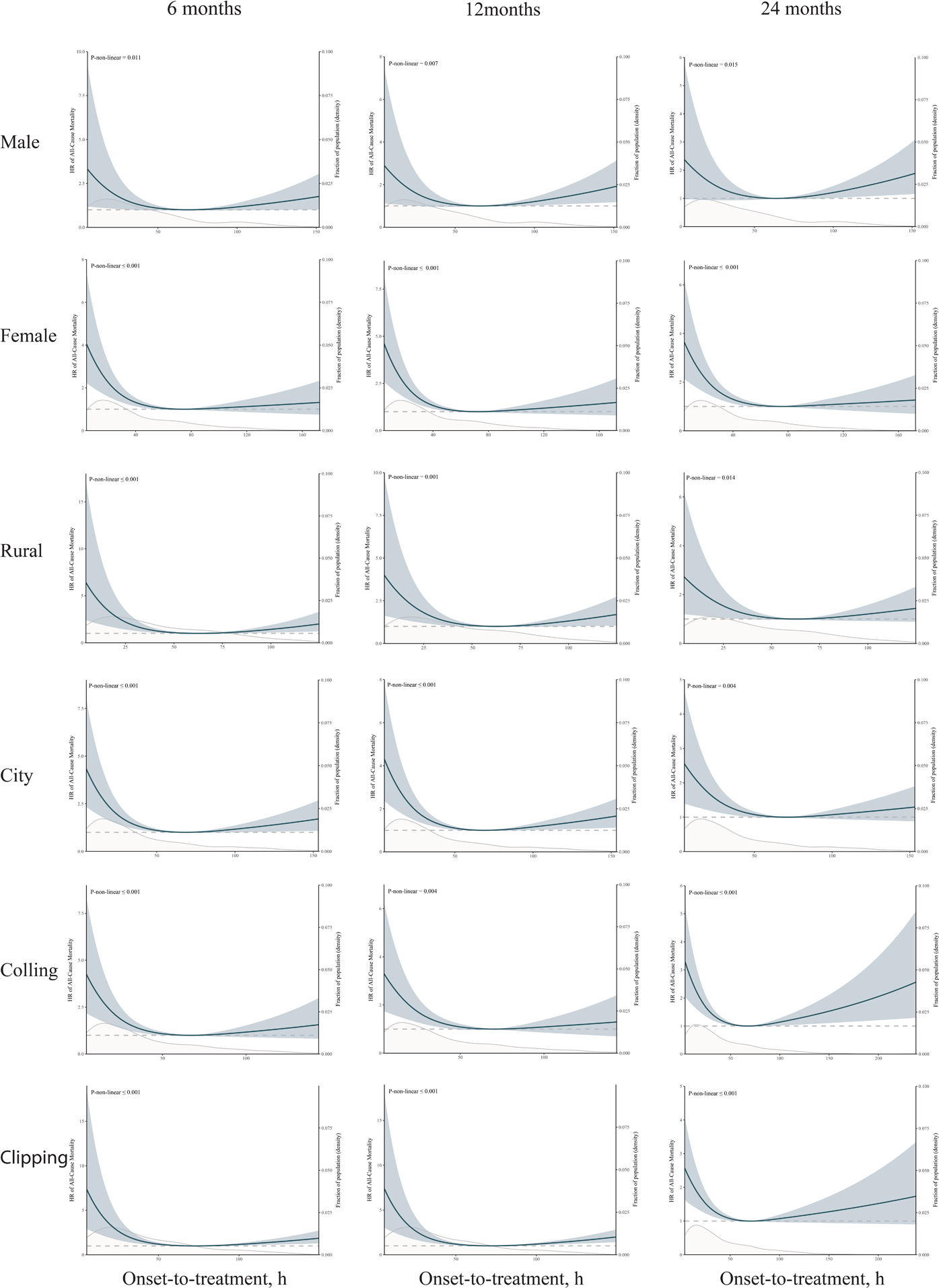

## Discussion

Among patients with aSAH undergoing surgery in northern China, wait time was associated with 2-year mortality and other complications. This is the first study, to our knowledge, to characterize the nonlinear association between onset-to-treatment time for elderly aSAH patients (age ≥70 years) and 24-month survival and complications. Onset-to-treatment time presented a U-shaped association with all-cause mortality among elderly people in China. Specifically, early or delayed onset-to-treatment time predicted a higher risk of mortality. After we explored this association in different subgroups of participants, the results further supported the U-shaped association between onset-to-treatment time and mortality. Our analyses suggest that the optimal time window for receiving aneurysmal SAH treatment is approximately up to 68 hours postsymptom onset.

Significant U-shaped associations of onset-to-treatment time with 24-month all-cause mortality were evident for elderly aSAH patients (age ≥70 years). Most observational studies have found that early and delayed onset-to-treatment time predicted mortality risk, ^3,12^ although some observational studies reported no relation of onset-to-treatment time with mortality.^22^ A 12-month follow-up among 482 participants in Australia showed that the correlation between onset-to-treatment time and mortality was nonlinear.^12^ However, none of those findings of a U-shaped association were based on Cox models with restricted cubic splines, which were able to examine a nonparametric (nonlinear or linear) association of onset-to-treatment time with mortality among elderly aSAH patients.

Performing surgery from 49 to 68 hours after symptom onset represents a substantial change in practice because 87.7% of the patients in this study did not receive surgery within this timeframe, highlighting the potential need to improve timely access to care for patients presenting to the hospital with aSAH symptoms. The optimal time window for receiving neurosurgical and endovascular treatment is approximately up to 68 hours after onset. Advanced microsurgery and endovascular treatment could offset the side effects of early surgery and achieve better outcomes than delayed surgery. Many studies have suggested that early surgery (treatment <3 days) is superior to delayed surgery (treatment >3 days) in reducing a poor outcome and death rate. The timing (within 1 day, 2 days, or 3 days) of definitive management of acutely ruptured intracranial aneurysms (neurosurgical or endovascular treatment) has been the subject of considerable debate. In the ISAT, the mean time to treatment was 1.1 days for endovascular coil embolization vs. 1.8 days for surgery.^23,24^ Large studies during the 1980s indicated that 4–10 days after aSAH was a particularly precarious time for surgery when cerebral vasospasm and cerebral ischemia may be the most active.^25^ The European Stroke Organization Guidelines recommend the timing of intervention: aneurysms should be treated as early as logistically and technically possible to reduce the risk of rebleeding; if possible, interventions should be aimed to intervene at least within 72 h after onset of first symptoms.^9^

The timing of ultra-early surgery (treatment <1 day) for aSAH remains controversial. In a study, the risk of death at 12 months was lowest when treatment occurred up to 12.25 hours after aSAH symptom onset but then increased from 12.25 hours onward.^12^ The main reason for ultra-early surgery is to avoid the known risks of rebleeding. In patients who survive the initial hemorrhage, rebleeding is also the major cause of morbidity and poor outcome.^26^ Multiple studies have shown that the incidence of rebleeding is maximal in the first 24 hours, with rates of 4.1% to 17.3% reported; therefore, a protocol of 24-hour emergency treatment as a way to reduce morbidity and mortality due to aneurysmal rebleeding is advocated. Recent reports focusing on hourly changes (the peak was within 2 hours, 3 hours, or 6 hours) in the rate of rebleeding have indicated more consistent results.^27^ The rates of the patients admitted within 2 hours, 3 hours, or 6 hours after the initial hemorrhage were lower, thus preventing rebleeding from occurring in the ultra-early period after the initial SAH. A single-center observational study reported that the implementation of 24-hour ultra-early aneurysm repair would have only resulted in a 0.3% reduction in the incidence of rebleeding. ^20^

Age was probably a factor influencing the effect of ultra-early surgery. Ultra-early (<24 hours after subarachnoid hemorrhage) coiling of ruptured aneurysms was marginally associated with improved clinical outcomes compared to coiling at ≥24 hours in elderly patients.^28^ A meta-analysis on treatment <1 day compared with any later treatment included strictly selected patient subsets based on age on admission.^10^ No benefit of treatment <1 day was found when compared with treatment on day 2 or 3 post-SAH. The reasons impeding the application of ultra-early surgery included suboptimal operating conditions in emergency surgery, highly reactive brain tissue, and poor patient tolerance. Elderly patients with a higher frequency of comorbidities and poor tolerance to ultra-early surgery would probably have their operations postponed. On the other hand, advances in critical care management, such as strict blood pressure control, cerebral vasospasm prevention, reversal of coagulopathies, pain management, nausea and emesis prevention, and improved neurocritical care, may reduce the rate of aneurysmal rebleeding prior to surgery.

### Strengths and limitations

This study has several strengths. First, our study had a large sample size, with consecutive patients across 12 centers, to assess the role of onset-to-treatment time in all-cause mortality and complications among elderly patients from northern Chinese provinces. This large sample allowed robust conclusions in establishing a time-to-surgery threshold for elderly aSAH patients before the risk of mortality increased. Second, rather than arbitrarily dividing patients into early and delayed surgery groups,^3,5,10^ exact time-to-surgery data (in hours) and Cox models with spline regression were used to empirically define a threshold for increased risk. Third, we adjusted for several confounders with an effect on mortality risk.

This study also has several limitations. First, the results of this study are subject to the well-known methodological limitations of retrospectively analyzed cohorts. Given the importance of this evidence in establishing clinical practice guidelines for SAH management, an adequately powered RCT is needed. Second, seizure events would be expected to occur more often in the delayed surgery group due to treatment delay. Because we did not have access to individual patient seizure data, it was impossible to determine whether onset-to-treatment time would have affected the seizure event. Therefore, we did not feel seizures were a valid outcome measure for this study. Third, the timing of treatment may be influenced by a number of factors that may also affect outcomes.^10^ Outside hospital transfer and poor clinical condition on admission are critical factors that influence onset-to-treatment time and clinical outcomes, and attempts were made in this study to address potential confounding by these variables through sensitivity analyses. On the other hand, for medically complex elderly patients with a higher frequency of comorbidities, several analyses were performed to mitigate the influence of confounding factors. Comparisons between middle-time surgery and early-time or late-time surgery groups after matching were balanced across more than 20 covariates. However, the finding that increased onset-to-treatment time was associated with increased risk may still have been influenced by unmeasured factors. Fourth, patients with nonoperative aSAH were not included because it was not possible to distinguish between those who died while awaiting surgery and those for whom nonoperative treatment was indicated. However, patients who experienced a major complication while waiting for surgery were excluded from the cohort, leading to more conservative estimates of the effect of waiting for surgery.

## Conclusions

In this study, among elderly aSAH patients in northern Chinese provinces, onset-to-treatment time showed a U-shaped association with 24-month all-cause mortality. Early surgery was superior to delayed surgery in reducing the death rate. Elderly patients with poor tolerance to ultra-early surgery probably had their operations postponed. A wait time of 68 hours may represent a threshold that defines a higher risk.

## Data Availability

The data that support the findings of this study are available from the corresponding author upon reasonable request.

